# Pilot Feasibility Study of a Community Health Worker-delivered Tobacco Cessation Intervention During Pregnancy: The Smoke-Free HOPE Randomized Controlled Trial

**DOI:** 10.64898/2026.09.29.26364272

**Authors:** Anne Berit Petersen, Berenice Rosas, Juan Carlos Belliard, Janice Tsoh, Sharlene Gozalians, Kelsey Quema, Bryan Oshiro, Ruofan Yao, Pramil N. Singh

**Affiliations:** Loma Linda University School of Nursing, Loma Linda, CA 92350 USA; Loma Linda University Cancer Center, Transdisciplinary Tobacco Research Program, Loma Linda, CA 92350 USA; Loma Linda University School of Public Health, Loma Linda, CA 92350 USA; Loma Linda University Health, Institute for Community Partnerships, Loma Linda, CA 92350; University of California San Francisco, San Francisco, CA 94143 USA; Loma Linda University School of Medicine, Loma Linda, CA 92350 USA; Riverside University Health System, Moreno Valley, CA 92555 USA; Loma Linda University Children’s Hospital, Department of OB/GYN - Maternal Fetal Medicine, Loma Linda, CA 92350

## Abstract

**Introduction:** Tobacco use during pregnancy increases risks for adverse birth outcomes. In the US, tobacco use during pregnancy is associated with several poverty-related social determinants of health (fewer years of education, housing and food insecurity, substance abuse) that also contribute to adverse birth outcomes. In the Smoke-free Household and Parent (Smoke-free HOPE) pilot feasibility study, we designed and tested a theory-based intervention (socio-ecological framework) that leverages our group’s ten years of experience enrolling and evaluating the largest maternal smoking cessation program in San Bernardino County. The Smoke-free HOPE intervention is delivered by community health workers (CHW) and integrates tobacco cessation support into the other support services that are typically offered during perinatal CHW visits. To our knowledge, this is the first US-based study to test a CHW-delivered prenatal tobacco cessation intervention.

**Methods:** The Smoke-free HOPE intervention is a two-arm RCT that randomizes pregnant adults (n=32) who use tobacco products (combustible, e-cigarettes/vapes) to one of two CHW-delivered interventions: brief behavioral advice or patient navigation to community-based tobacco cessation services (i.e., state quitline). Both intervention arms were delivered during up to eight CHW visits (home, telehealth). Primary feasibility and acceptability outcomes are reported.

**Results:** At baseline, we found that the predominant form of habitual tobacco use was e-cigarettes (53%) and that co-use of cannabis occurred in 69%. Two participants switched from cigarettes/cigars (“blunts”) to e-cigarettes during the trial. For feasibility, we met our recruitment and retention progression criteria by achieving the following: a county-based enrollment of four consenting participants per month, 87% initiating CHW support (i.e., dropout rate of <13%), and 66% completing all eight CHW visits. For post-intervention acceptability, 91% were very satisfied with the program, and 100% would recommend it to their peers. Post-intervention key informant interviews revealed that participants valued the trust, flexibility, and nonjudgmental support of CHW visits. Seventy-five percent [95% CI = 60% to 90%] set a quit date, and 50% [95% CI = 33% to 67%] achieved 30-day point-prevalence abstinence from nicotine.

**Conclusions:** We have documented the feasibility and acceptability of a CHW-delivered intervention to provide cessation support for all major tobacco products used by pregnant adults in our community. Scaling the intervention to multiple sites will help us measure its efficacy.

**Implications:** This pilot study provides “proof of concept” findings of the feasibility and acceptability of a CHW-delivered intervention for cessation of prenatal tobacco use. Smoke-free HOPE is the first US-based RCT to test a CHW model for tobacco cessation during pregnancy. At our San Bernardino County study site, we found that CHW home visits during 2024-2026 substantially decreased the dropout rate (36% to 13%), compared with our previous program (2012-2019) with perinatal health educators and a classroom setting. Also noteworthy is that Smoke-free HOPE provided cessation support to more than half of our trial participants who enrolled in order to quit e-cigarettes/vapes. Post-program interviews with half of the trial participants found that CHW visits became a source of trusted, flexible, and nonjudgmental support for their quit attempt.

## 1. Introduction

Tobacco use continues to be the most common form of substance use during pregnancy (1, 2). The 2024 U.S. Surgeon General’s Report reaffirmed that tobacco use during pregnancy contributes to adverse birth outcomes, including preterm birth, low birth weight, fetal growth restriction, and infant mortality (3, 4). In addition to combustible tobacco, recent evidence links non-combustible nicotine products, including e-cigarettes (5), chewing tobacco/snus (6), betel quid (7), and iqmik (8) to adverse birth outcomes, particularly low birth weight, preterm birth, and fetal growth restriction. Consequently, professional and public health organizations, including ACOG, CDC, USPSTF, and WHO, have emphasized that nicotine-containing products, including e-cigarettes/vapes, are not considered safe during pregnancy and that pregnant patients should be screened and supported in cessation efforts (9–12).

Given the expanding range of nicotine products used during pregnancy in the US, effective cessation strategies must address both combustible and non-combustible nicotine products and account for increasingly complex patterns of dual and poly-product use. Despite this need, evidence-based cessation interventions for non-combustible nicotine products remain limited. Notably, in the US, there are currently no FDA-approved pharmacologic or behavioral interventions specifically indicated for e-cigarette cessation, and evidence-based guidance for helping individuals quit vaping remains sparse (10, 13).

Prenatal tobacco use disproportionately affects populations experiencing socioeconomic and structural disadvantages. In the United States, higher prevalence of prenatal tobacco use is consistently associated with lower educational attainment, lower income, Medicaid enrollment, rural residence, depression, and certain racial and ethnic groups, including American Indian/Alaska Native, Black/African American, and White populations (14–16). Although non-pharmacologic interventions can improve cessation during pregnancy (17), effectiveness may be lower among individuals facing socioeconomic disadvantage, heavier dependence, and pro-smoking social environments. This highlights the need for more intensive, tailored approaches for high-risk communities (18).

In San Bernardino County – a large California county with one of the highest maternal morbidity rates (19, 20) – our group developed the Comprehensive Tobacco Treatment Program (CTTP) – the largest state-funded (First5CA.gov) maternal tobacco cessation program. The CTTP consisted of a behavioral smoking cessation program tailored to pregnant women and delivered by health educators in a small-group, in-person format (21). Although CTTP demonstrated the efficacy (i.e., 40% abstinence rate, significant decrease in low birthweight for those who quit) of delivering pregnancy-specific tobacco cessation, our 2012-2019 enrollment findings also highlighted challenges such as a high rate of dropouts and no consideration of the modern tobacco product exposome (new generation of e-cigarettes/vape products) (21). Also, by race/ethnicity, the groups with the highest state and national prevalence of tobacco use during pregnancy (African American/Black, Native American, White) also had the highest relapse rate after enrollment in CTTP (22).

These findings informed our intervention mapping process and the design of the Smoke-free HOPE (<u>Smoke-free H</u>ouseh<u>O</u>ld and <u>P</u>ar<u>E</u>nt) program, which delivers behavioral models of prenatal tobacco cessation through Community Health Worker (CHW) home visits (telehealth, in-person) intended to increase program retention and nicotine abstinence. We specifically hypothesize that including tobacco cessation support into the other support services (23–28) that are typically offered during perinatal CHW visits will be feasible, acceptable, and ultimately scalable. For this study, we define CHWs using the American Public Health Association definition, encompassing a distinct frontline public health workforce with defined roles and competencies and, in our setting, competency-based training and certification requirements (29). Thus, our CHW model is not intended to be directly comparable to findings from peer, lay, or village health workers.

In this report, we present the pilot feasibility findings of the Smoke-Free HOPE trial. To our knowledge, Smoke-Free HOPE is the first U.S.-based RCT to test the feasibility of using trained CHWs to deliver a pregnancy-tailored tobacco cessation program during prenatal care (30). The overall aim of the Smoke-Free HOPE trial was to assess the feasibility and acceptability of two behavioral models of CHW-delivered behavioral smoking cessation (brief advice through SCRIPT™ (Smoking Cessation and Reduction in Pregnancy Treatment)) (31) and patient navigation) tailored to pregnant individuals. The specific aims of Smoke-Free HOPE are: 1) To examine the feasibility of implementing CHW-delivered tobacco cessation support during prenatal care by measuring rates of recruitment and retention, and 2) To examine the acceptability of CHW-delivered tobacco/nicotine cessation support among pregnant participants. This report presents Aim 1 findings and Aim 2 acceptability findings from participants’ perspectives.

## 2. METHODS

### 2.1 Study Design and Oversight

Smoke-Free HOPE was a two-arm RCT to assess the feasibility and acceptability of two CHW-delivered tobacco cessation interventions during pregnancy for pregnant individuals using tobacco or nicotine products. The study included a qualitative component in post-intervention data collection to assess acceptability and identify opportunities to refine the intervention. The study protocol was previously reported (30). The Loma Linda University Institutional Review Board (IRB #5220112) approved the study, and all participants provided written informed consent. The trial was registered at ClinicalTrials.gov (NCT06438549) (32) and is reported in accordance with the CONSORT extension for randomized pilot and feasibility trials (32).

### 2.2 Setting

Participants were recruited from the Loma Linda University Health (LLUH) Maternal and Fetal Medicine clinic and community referral networks in San Bernardino County (i.e., federally qualified health centers, prenatal care clinics, and community-based organizations focused on maternal health) – a county-based patient catchment area of over 4 million (21).

### 2.3 Participants

#### 2.3.1 Eligibility Criteria

Eligible participants were pregnant adults aged 18 years or older who were current users of combustible or non-combustible tobacco/nicotine products or who had recently quit during pregnancy but were considered at risk for relapse. Eligibility included any use of combustible cigarettes, e-cigarettes/vapes, and other nicotine-delivery products (chewing, snus, oral nicotine pouch). Participants were also required to be willing to quit and set a quit date within two weeks of enrollment. Individuals were excluded if they had severe mental health conditions that precluded informed consent or participation, were on nicotine replacement therapy (NRT), or were currently enrolled in another cessation program.

#### 2.3.2 Recruitment, Consent and Enrollment

Participants were recruited using a multimodal strategy that included (1) provider referral during prenatal care visits, (2) electronic health record–based identification of potentially eligible patients, and (3) community outreach, including self-referral and referrals from community-based organizations. Study staff conducted initial eligibility screening using a two-step consent process. At the initial contact, study staff requested verbal permission to ask brief screening questions to determine preliminary eligibility. Individuals who appeared eligible after this prescreen were invited to complete the full study consent process before any baseline assessment or intervention activities. This included written informed consent, permission to access and use relevant health information, and review of participant rights. We enrolled participants only after clinical review confirmed that they met all eligibility criteria (30).

### 2.4 Randomization and Blinding

Participants were randomized in a 1:1 ratio to one of two intervention arms using a block randomization schedule generated in SAS software (Cary, NC). The study program coordinator managed allocation. Because the interventions were behavioral in nature, participants and CHWs were not blinded to treatment assignments. To minimize potential bias, we used standardized outcome assessment procedures, and analytic approaches emphasized objective and descriptive measures where feasible.

### 2.5 Interventions

#### 2.5.1 Intervention overview

Both intervention arms consisted of up to eight weekly sessions delivered by trained CHWs through home visits or telehealth encounters. Sessions were embedded within a standard CHW visit structure and included check-ins, intervention-specific tobacco cessation support, screening for health-related social needs, referral or navigation support as indicated, and visit documentation (33). CHWs used participant-centered motivational communication approaches across both arms. Missed sessions were rescheduled when possible, and weekly outreach continued during the intervention period unless the participant explicitly declined further intervention contact or formally withdrew. We assessed intervention exposure and completion using predefined engagement metrics, as described in the Feasibility Outcomes section.

#### 2.5.2 CHW-Brief Advice (CHW-BA)

The CHW-BA intervention consisted of CHW-delivered behavioral cessation support adapted from the evidence-based SCRIPT™ program materials (31, 34). The intervention emphasized motivation, self-efficacy, quit planning, practical cessation skills, supporting quit planning, and relapse prevention. Content was adapted to address both combustible and non-combustible nicotine products, including e-cigarettes and dual-use patterns. Table 1 summarizes the core intervention elements.

#### 2.5.3 CHW-Patient Navigation (CHW-PN)

The CHW-PN intervention consisted of structured, CHW-delivered navigation support designed to connect participants with evidence-based cessation resources and sustain engagement over time. Using a curated list, CHWs help participants identify appropriate cessation services and support, including quitlines, digital cessation tools, and community-based resources; facilitate enrollment when needed; and provide follow-up, troubleshooting, and re-engagement support throughout the intervention period.

#### 2.5.4 CHW Interventionists and Training

Both interventionists were formally trained CHWs with prior field experience; one completed the 6-month Loma Linda University Health San Manuel Gateway College CHW/Promotores Program, and the other entered the study with prior CHW certification and experience. Before intervention delivery, both completed a 3-day study-specific training covering tobacco cessation in pregnancy, motivational interviewing, and arm-specific procedures for either SCRIPT™-based brief counseling or patient navigation; full training details are reported in the study protocol (30).

#### 2.5.5 Intervention Process Evaluation

CHWs documented intervention delivery using structured activity logs that captured session completion, duration, modality, intervention content delivered, referrals or navigation activities, and participant engagement. Standardized protocols, ongoing supervision, and CHW debriefing ensured consistent intervention delivery across study arms and documented adaptations, delivery challenges, and contextual factors relevant to implementation.

### 2.6 Outcomes

#### 2.6.1 Feasibility Outcomes

The primary feasibility outcomes included recruitment and retention. We assessed recruitment by monthly enrollment rate. We defined retention using indices of CHW visit completion. We considered participants who did not initiate any CHW visits after randomization as dropouts; we also reported retention across all eight CHW visits.

#### 2.6.2 Acceptability Outcomes

We assessed acceptability using participant surveys and qualitative interviews. Survey measures assessed participant satisfaction, perceived usefulness of the intervention, perceived support from CHWs, and participant perceptions of program characteristics associated with engagement and behavior change using adapted items from the Critical Factors Survey (30, 35). Qualitative data further assessed participant experiences, perceived benefits and burden, and recommendations for intervention refinement.

#### 2.6.3 Exploratory Tobacco/Nicotine Outcomes

Exploratory tobacco and nicotine outcomes included setting a quit date and self-reported 7-day point-prevalence abstinence from all tobacco products at 1 month. The study was not powered to detect statistically significant differences between study arms for these outcomes.

### 2.7 Data Collection and Measures

CHWs documented intervention activities, tobacco and nicotine use, and quit status at each intervention session using standardized activity logs. Research staff administered study assessments separately via telehealth at baseline and at 1, 3, and 6 months after the participant’s quit date. These assessments included self-reported tobacco and nicotine use and other study outcomes, as specified in the study protocol (30). Participants who completed all 8 CHW intervention visits were also invited to complete the postintervention acceptability survey and interview. Clinical and demographic information was obtained through participant self-report and electronic health record review.

### 2.8 Qualitative Methods

All participants who completed the eight-session CHW intervention were eligible and invited by the program coordinator (PC) via telephone, email, and text to complete a post-program semi-structured interview. The qualitative component is reported in accordance with the Consolidated Criteria for Reporting Qualitative Research (COREQ) checklist (36) (Supplemental File 1). The pilot-tested interview guide, informed by COM-B constructs (Supplemental File 2), explored intervention experiences, perceptions of CHW support, barriers and facilitators to engagement, smoking and vaping cessation, and recommendations for program improvement. Interviews were conducted via secure videoconference (mean 50 minutes; range 33–65), audio-recorded, and professionally transcribed verbatim. We recorded field notes; transcripts were not returned for review; no repeat interviews occurred; and no other adults were present, although infants were occasionally present. Because we invited all eligible completers, interview accrual was determined by the trial sample and participant uptake rather than data saturation. Two female study investigators (ABP and BR) conducted interviews and had qualitative research or study-specific interviewing training. Neither knew participants before enrollment, although BR had study-related contact; participants knew the interviewers’ roles in the study. We managed de-identified transcripts in NVivo (37). Two investigators (SG and ABP) analyzed the data using a hybrid deductive-inductive thematic analysis (38, 39). SG initiated coding using an a priori framework derived from the study objectives, interview guide, and implementation constructs related to feasibility, acceptability, appropriateness, engagement, participant burden, and program refinement. Additional inductive codes captured concepts, experiences, and mechanisms emerging from participant narratives. Through iterative cycles of familiarization, memo writing, descriptive and focused coding, investigators refined code definitions, consolidated related concepts, and developed higher-order thematic categories. Supplemental File 3 provides the final coding tree, and analytic memos document coding decisions, evolving interpretations, thematic relationships, and convergent and divergent experiences.

Cross-case comparisons examined similarities and differences across participants and intervention arms, with attention to acceptability, appropriateness during pregnancy, flexibility, burden, engagement, CHW relationships, cessation experiences, and recommendations for refinement. We supported rigor through an audit trail, constant comparison of recurring and divergent cases, and iterative team discussion of interpretations, code definitions, and thematic relationships. These discussions challenged assumptions and explored alternative explanations to strengthen reflexivity and analytic depth rather than establish coder agreement or inter-rater reliability. Themes were generated through iterative synthesis of coded data, and representative quotations were selected to illustrate participant experiences while preserving confidentiality.

### 2.9 Statistical Analysis

Analyses focused primarily on descriptive evaluation of feasibility and acceptability outcomes. For quantitative outcomes (recruitment, retention, and acceptability), we reported proportions with 95% CI; for comparisons of proportions, we reported the chi-squared test for independence (40, 41). When cell counts were low, we used non-parametric procedures (Fisher’s exact test, non-parametric bootstrapped confidence intervals) (40, 41). For continuous variables in descriptive analyses and on acceptability surveys, we reported means and 95% confidence intervals. Comparisons were made by independent samples t-tests. Similarly, for small sample sizes, we used non-parametric methods (Wilcoxon’s Rank Test, non-parametric bootstrapped confidence intervals). All analyses were conducted on SAS version 9.4 (Cary, NC).

### 2.10 Integration of Quantitative and Qualitative Data

Quantitative and qualitative data were analyzed separately and integrated during interpretation using a joint display (42). We compared feasibility and acceptability indicators with corresponding participant interview themes to identify convergence, complementarity, and divergence. Broader analyses of multilevel implementation barriers, facilitators, and CHW intervention processes will be presented in another report.

## 3. Results

### 3.1 Smoke-free HOPE Participant Flow and Characteristics

Figure 1 shows participant flow through screening, enrollment, randomization, follow-up, and analysis. We randomized 32 participants to CHW-BA (n=17) or CHW-PN (n=15). Table 2 provides the demographic characteristics of the Smoke-Free HOPE trial participants. At baseline, the mean age was 31 years; 44% of subjects were Black/African American; 6% had a college or higher education; 63% were unemployed; 22% were married; and 70% participated in at least one public assistance program. The predominant form of tobacco used was e-cigarettes (53%), followed by cigarettes (41%). None of the participants used e-cigarettes as a cessation aid for combustible tobacco. None of the participants reported use of oral nicotine pouches (i.e., habitual or as a cessation aid). We also found that 69% of subjects had used cannabis in the last year.

### 3.2 RCT Feasibility Findings

After activating all recruitment methods (electronic medical record, provider referral, community referral), we recruited 4 subjects per month within San Bernardino County health systems. This rate meets our published progression criteria (30) and supports the feasibility of using our San Bernardino catchment area as an enrollment site in a multi-site trial with a 3–4-year enrollment period. Notably, 87% of participants initiated CHW sessions, and 13% were considered dropouts due to not initiating CHW sessions (Figure 1). Notably, none of the subjects who dropped out did so by formally declining the intervention by contacting study staff. Sixty-six percent completed all eight CHW visits. We note that the CHW visits incorporated the intervention (i.e., two models of CHW-delivered tobacco cessation support) into standard-of-care CHW visits that also addressed social determinants of health (SDOH). Among participants who received CHW support, 96% also received support for SDOH-related needs during one or more of their CHW visits. Supplemental File 4 presents the distribution of SDOH-related needs addressed during the trial.

### 3.2 RCT exploratory findings for Nicotine Abstinence

Our study was not powered to compare point prevalence abstinence from nicotine between the two CHW-led intervention arms (CHW-BA, CHW-PN). For all trial participants (n=32), we found that 75% [95% CI = 60% to 90%] set a quit date, and 50% [95% CI = 33% to 67%] achieved 30-day point prevalence abstinence from nicotine. Among all e-cigarette users seeking cessation support at baseline, 76% [95% CI = 56% to 97%] set a quit date, and 53% [95% CI = 29% to 77%] achieved 30-day point prevalence abstinence from nicotine. Arm-specific findings showed no significant differences between arms in achieving 30-day point prevalence abstinence from nicotine (CHW-BA, 47% [95% CI = 23% to 71%]; CHW-PN, 60% [95% CI = 35% to 84%], p=0.46).

### 3.3 Mixed Methods Acceptability Findings

Our mixed-methods acceptability findings come from post-intervention surveys completed by 16 subjects and key informant interviews completed by 16 subjects.

#### 3.3.1 Quantitative Findings: Survey data

Table 3 presents the survey findings and indicates that most participants were very satisfied with the overall program (91%) and the support received (91%), and 100% reported that they would ‘definitely’ recommend Smoke-Free HOPE to their peers. Respondents rated CHW communication highly (mean 9.7/10; range 7–10). Among CHW support characteristics, reliability and consistency and celebrating small achievements were most consistently reported as fully achieved (91% each), followed by a nonjudgmental approach and clear boundaries (90% each). All participants who evaluated CHW-provided resources and Kick It California resources rated them as very helpful. (<u>Kick It California</u> offers free cessation support through Quit Coaches, smoking- and vaping-specific text programs, a mobile app, and self-help materials).

#### 3.3.2 Qualitative Findings: Post-program Participant Interviews

Of 22 eligible participants, 16 completed post-program qualitative interviews. No participants formally declined; the six nonparticipants did not respond to telephone or text invitations. Among 16 interview participants (mean age 30 years; range, 20–37), 37.5% were African American/Black, 31.3% Latino/Hispanic, 25.0% mixed race/ethnicity, and 6.3% White; 62.5% were in CHW-BA and 37.5% in CHW-PN. Half primarily/exclusively vaped; the remainder reported cigarette-only (12.5%), dual cigarette/e-cigarette (12.5%), or other co-use (25.0%). Interviews contextualized participant engagement and acceptability among completers and identified broad priorities for intervention refinement. Four themes relevant to these outcomes are presented, followed by arm-specific feedback on intervention resources. Illustrative quotations are identified by participant ID, with divergent experiences noted where relevant.

##### 3.3.2.1 Relational, flexible, and nonjudgmental CHW support shaped engagement and acceptability

Participants consistently identified the CHW relationship as a valued component of the intervention, describing CHWs as sources of encouragement, accountability, emotional support, and practical guidance. Several participants contrasted CHW interactions with prior healthcare experiences in which they had felt judged or stigmatized for nicotine use during pregnancy. As one participant explained, “She was the only one I felt comfortable talking to” [Participant ID: SFH-12],

Participants valued continued CHW support when they relapsed, missed goals, or struggled to reduce nicotine use, particularly while managing parenting, financial, relationship, and other competing demands, as one participant reflected, “She made me feel like I wasn’t failing” [SFH-05]. Flexibility also reduced participation burden. Scheduling accommodations, telephone and text communication, and virtual visits helped participants remain involved despite competing demands. One participant noted, “They worked around my schedule and checked in when I needed it ” [SFH-21]. Among interviewed completers, trust, flexibility, accountability, and nonjudgmental support were closely associated with continued engagement.

##### 3.3.2.2 Pregnancy strengthened motivation, while participants valued support that acknowledged the nonlinear nature of cessation

Pregnancy was a strong source of motivation to quit or reduce tobacco and nicotine use. Participants described concern for fetal and infant health, breastfeeding, secondhand smoke exposure, and becoming healthier parents; for some, pregnancy prompted their first serious cessation attempt. One participant explained, “I wanted to stop because I didn’t want to hurt my baby” [SFH-28]. At the same time, participants described cessation as difficult and often nonlinear, with setbacks occurring despite strong intentions to quit.

Participants particularly valued CHW support that acknowledged these challenges rather than focusing exclusively on immediate abstinence. They appreciated encouragement following setbacks and support that recognized gradual progress and repeated quit attempts. As one participant explained, “It helped knowing someone understood how hard it was instead of just telling me to stop” [SFH-23]. These accounts suggest that intervention acceptability was partly related to CHW support that accommodated the complexity and nonlinear nature of behavior change during pregnancy.

##### 3.3.2.3 Stress, withdrawal, vaping accessibility, and social environments complicated cessation efforts

Participants described emotional distress and nicotine dependence as important challenges during cessation efforts. Smoking and vaping were frequently used to manage stress, anxiety, loneliness, parenting demands, grief, and relationship conflict, making nicotine use more than a habitual behavior for some participants. One participant explained, “When life got overwhelming, that’s when I relapsed” [SFH-20]. Cravings and withdrawal further complicated quit attempts, particularly emotional symptoms such as irritability, anxiety, frustration, crying, and feeling overwhelmed.

Participants who vaped described additional challenges related to convenience and constant availability. Vaping could occur indoors, in vehicles, while parenting, and during routine activities, making use less bounded than cigarette smoking. As one participant stated, “I could vape basically anywhere” [SFH-10]. Some participants also described perceptions of vaping as less harmful than cigarettes. Social environments further reinforced nicotine use when partners, family members, or peers continued to smoke or vape, creating repeated exposure to cues and normalizing use. In less common accounts, participants also described cannabis or alcohol use as influencing their tobacco or e-cigarette use, illustrating the potential role of co-use in cessation experiences. Together, these accounts show how emotional coping, cravings and withdrawal, product accessibility, and social context complicated cessation efforts despite strong motivation to quit.

##### 3.3.2.4 Participant experiences identified opportunities for intervention refinement

Participants identified several priorities for strengthening future iterations of the intervention, including more vaping-specific cessation support, greater attention to relapse prevention and emotional coping, and continuation of support into the postpartum period. Participants favored approaches that normalized setbacks and recognized reduction and repeated quit attempts as part of the cessation process. As one participant explained, “As one participant explained, “I hit the vape the two times I slipped up, [and] she was never mad” [SFH-005].

The postpartum period was identified as a particular opportunity for continued relapse-prevention support. Participants noted that formal intervention support ended as parenting demands, sleep disruption, and other stressors could increase. One participant suggested, “So maybe keep the support going… at least a week or two after the baby is born… because that’s when the stress is going to be mostly there… and you might want to pick up and use again” [SFH-08]. Participants also noted the influence of partners and household members who used nicotine, suggesting potential value in considering family or household context in future intervention refinement.

##### 3.3.2.5 Arm-specific feedback on intervention resources

Experiences with arm-specific resources identified opportunities for refinement. In the CHW brief-advice arm, participants described the SCRIPT™ booklet (38) as a useful structure for discussing motivation, quit planning, and coping. CHWs adapted the cigarette-focused material for participants who vaped and supplemented it with vaping cessation resources. Although helpful during CHW-guided sessions, the booklet was not consistently revisited between visits and was sometimes misplaced, suggesting a need for more accessible, vaping-specific resources.

In the CHW patient-navigation arm, participants described navigation as extending beyond referral. CHWs helped overcome practical and technological barriers to using Kick It California resources, including the app and cessation counseling, and provided ongoing follow-up and accountability. As one participant explained, “She encouraged me every time we met… ‘We’re going to utilize it today.’… So, we would call together” [SFH-016]. Motivational messages, progress tracking, quit coaching, and chat support also provided support between CHW contacts and, for some, beyond the intervention. Together, these accounts suggest active facilitation, troubleshooting, and follow-up supported engagement with cessation resources.

#### 3.3.3 Integration of Quantitative and Qualitative Findings

Table 4 presents the joint display integrating quantitative and qualitative findings. High acceptability converged with participant reports of trusted, flexible, accessible, and nonjudgmental CHW support. Qualitative findings also contextualized engagement and cessation indicators by highlighting setbacks, stress, withdrawal, product accessibility, and social influences. These findings informed priorities for vaping-specific support, postpartum relapse prevention, and active patient navigation through facilitation, troubleshooting, and follow-up.

## 6. Discussion

To our knowledge, Smoke-free HOPE is the first U.S.-based RCT to evaluate the use of certified CHWs (certificate-based training for clinic-based CHW practice) to deliver prenatal tobacco cessation. In the US, CHWs are increasingly integrated into healthcare systems – particularly federally qualified health centers – to improve social determinants of health and improve access to care in underserved populations (43, 44). Here, we report on the feasibility of adding tailored tobacco cessation support to the current CHW visit schedule during prenatal care.

Our findings from Smoke-free HOPE indicate that we met our published (30) feasibility criteria (recruitment, retention) and suggest that adding tobacco cessation support to the CHW role during prenatal care is scalable. Specifically, our recruitment methods (electronic medical record invitation, provider referral, community and self-referral) produced an enrollment of four consented subjects per month from the San Bernardino County study site. This supports the feasibility of including our study site in a future multisite trial with 3–4 years of enrollment. The dropout rate for the Smoke-free HOPE model of intervention delivery through CHW visits (13%) was also well below the progression criterion we set based on the dropout rate (36%) we achieved using perinatal health educators in a classroom setting in San Bernardino County (21).

In our multi-ethnic trial sample, acceptability was high as evidenced by 91% participant satisfaction with the intervention, 100% participant recommendation to peers, and a 9.7 out of 10 score for CHW communication effectiveness. Qualitative findings similarly emphasized trusted, flexible (home and telehealth visits), and nonjudgmental CHW relationships as important to engagement, including during relapse, and amidst competing life stressors. Our previous CTTP model for San Bernardino County (21, 22)used perinatal health educators that focused only on tobacco cessation, and we found that by race/ethnicity, the groups that had the highest prevalence of prenatal tobacco use (Alaskan Native/American Indian, Black/African American, White) also had the highest relapse rate from CTTP. We posit that the high acceptability of the Smoke-free HOPE CHW model may be because it was delivered as part of a more whole person care model with outreach further enhanced through CHW visits (home, telehealth). Notably, although we lost some participants to follow-up, none formally withdrew from the intervention by contacting CHWs or study staff.

Preliminary cessation outcomes were encouraging: 75% set a quit date, and 50% reported 30-day point-prevalence abstinence from nicotine. More than half enrolled seeking support to stop e-cigarette use, and this finding validated the advisement we received from our Community Advisory Board to expand the cessation program to e-cigarettes/vapes. Notably, participants attempting to quit e-cigarette/vapes achieved 53% 30-day abstinence from nicotine. Notably, two subjects did not achieve nicotine abstinence due to switching from combustible to e-cigarettes, and there was no other report of using other smokeless tobacco products (i.e., oral nicotine pouch, chewing tobacco, snus) during the trial for any reason.

### CHWs for delivery of Prenatal Tobacco Cessation

In Smoke-Free HOPE, the prenatal tobacco cessation intervention by certified CHWs as the primary interventionists (30) produced a 30-day point prevalence abstinence from nicotine from all forms of tobacco (50%) and among exclusive e-cigarettes/vape user (53%), which is higher than the national average for adults (all tobacco, e-cigarettes) reported by the PATH study (45, 46). Notably, the Smoke-free HOPE model added tailored prenatal tobacco cessation support to standard-of-care CHW support, which was also designed to address non-tobacco SDOH-related concerns (i.e., lower educational attainment, housing insecurity, food insecurity, access to care). Nearly all CHW visits in the trial (96%) involved significant time spent on other SDOH concerns unrelated to tobacco (See Supplemental File 4).

To date, the most comparable prenatal tobacco cessation interventions involving formally trained community health workers were conducted outside the US and used different models specifically designed to address high rates of prenatal tobacco use among Indigenous populations in New Zealand (47, 48) and Australia (35). In New Zealand, Moai CHWs (‘Aunties’) with formal training received study-specific training to recruit pregnant women who smoked and provide cessation support, including access to free NRT (48). In this single-arm feasibility study, 8 of 33 participants with available birth records (24%) were documented as nonsmoking at delivery. In contrast to Smoke-free HOPE, which used a prescriptive set of eight scheduled CHW visits, the “Aunties” CHW-delivered intervention required an initial cessation-support visit, with subsequent contacts determined by the individual Auntie.

In Australia, the ICAN QUIT and the subsequent SISTAQUIT trials used a culturally tailored ABCD framework (Ask/Assess, Brief Advice, Cessation Support, and Discuss the psychosocial context) to strengthen prenatal smoking cessation care for Aboriginal and Torres Strait Islander women (35, 49). ICAN QUIT was a pilot step-wedge cluster-randomized feasibility trial, followed by SISTAQUIT, a multicenter pragmatic cluster RCT. Aboriginal Health Workers received study-specific cessation training alongside GPs, nurses/midwives, and other health professionals and were not designated as the primary interventionists. Rather, the intervention used a broader service-capacity-building approach to build multidisciplinary capacity for culturally responsive behavioral cessation support and access to free oral NRT (49, 50). In ICAN QUIT, 13.6% (3/22) of participants achieved CO-verified 7-day point-prevalence abstinence at 12 weeks (35).

### E-cigarette use during Pregnancy: From Cessation Aid to Cessation Target

CDC and ACOG have published formal guidance recommending avoiding e-cigarette use during pregnancy (1, 11). Notably, more than half of Smoke-Free HOPE participants enrolled were attempting to quit e-cigarette use, highlighting a US-based need for maternal tobacco cessation programs to focus on combustible and non-combustible tobacco products – a feature already included in federal health guidance. Evidence to guide the design of an e-cigarette cessation program for pregnant adults is limited. In this context, our exploratory finding that participants using e-cigarettes achieved a 53% 30-day point prevalence abstinence from nicotine is noteworthy. These findings also align with our post-intervention qualitative findings identifying participants’ requests for more e-cigarette/vape-specific guidance. Participants’ accounts also suggested that some perceived vaping as particularly difficult to discontinue.

Several features of vaping may warrant specific attention in prenatal tobacco cessation interventions. E-cigarette use can involve high nicotine exposure, difficulty monitoring consumption, frequent or habitual use, rapid reinforcement, and strong social and environmental cues (51, 52). These factors may be particularly relevant during pregnancy, when nicotine metabolism is increased. Pregnancy-specific qualitative studies similarly identify device characteristics, habitual use, and social and environmental influences as important determinants of vaping behavior (53–55).

Our findings also raise the possibility that prenatal nicotine cessation may occur within the broader context of prevalent cannabis co-use, another substance for which CDC and ACOG recommend avoidance during pregnancy (56–58). In Smoke-free HOPE, 69% of participants reported past-year cannabis use at baseline, including one participant who reported blunt use at enrollment. These observations further support models that can also consider co-occurring substance use during prenatal tobacco cessation interventions.

### Implications for Tobacco Control

In a recent US Medicaid claims analysis, only 6.3% of pregnant beneficiaries with documented tobacco use received cessation treatment during pregnancy, despite coverage of evidence-based treatment (59). Moreover, even when clinicians make referrals, engagement remains limited. The systematic review by Bailey et al. (2023) found that uptake of external smoking cessation appointments following referral during pregnancy was low and concluded that evidence on strategies to improve engagement remains insufficient (60). Based on these data and Smoke-free HOPE findings, we posit that CHWs may help bridge this referral-to-engagement gap by actively connecting patients with quitlines, digital interventions, and community-based resources, identifying options that fit individual needs, and reinforcing engagement over time. In this context, findings from our patient navigation arm are noteworthy.

### Limitations

We enumerate several limitations of our pilot feasibility trial here. Sample size and design precluded a statistical comparison of the CHW-delivery models (Brief Advice, Patient Navigation) between the two arms, or a comparison with a control. Our abstinence findings are based on self-report. Future work in this area needs to include biochemical verification of abstinence both as a study outcome and a “biofeedback” based intervention component.

## Conclusions

In a US multi-ethnic trial sample, we have documented the feasibility and acceptability of including prenatal tobacco cessation support into the “whole person care” workload of trained CHWs serving pregnant populations. Larger multisite trials are needed to measure the efficacy of the CHW-delivered interventions tested in the Smoke-free HOPE trial.

## Supporting information

Supplemental File 1. COREQ Checklist

Supplemental File 2. Interview Guide

Supplemental File 3. Coding Tree

Supplemental File 4. SDOH needs reported

## Ethics approval and consent to participate

The Loma Linda University Health Institutional Review Board granted ethical approval (Approval No. 5220112).

## Availability of data and materials

The data supporting this research are available from the authors on reasonable request.

## Author Contributions (CRediT)

**ABP:** Conceptualization, Methodology, Formal analysis, Funding acquisition, Project administration, Supervision, Writing – original draft, Writing – review & editing.

**PNS:** Conceptualization, Methodology, Formal analysis, Writing – original draft, Writing – review & editing.

**BO:** Methodology, Investigation, Writing – review & editing.

**JT:** Methodology, Writing – review & editing.

**JCB:** Methodology, Writing – review & editing.

**BR:** Investigation, Data curation, Formal analysis, Writing – review & editing.

**SG:** Formal analysis, Writing – original draft, Writing – review & editing.

**KQ:** Formal analysis, Writing – original draft, Writing – review & editing.

**RY:** Investigation, Writing – review & editing.

All authors contributed to interpretation of the findings and approved the final version of the manuscript.

## Acknowledgements

We thank the participants in Smoke-Free HOPE for their time, experiences, and contributions to this study. We also thank the Smoke-Free HOPE Community Advisory Board members for their sustained guidance and partnership throughout the study’s development and implementation. We thank the leadership and clinical team at Loma Linda University Health Maternal–Fetal Medicine for supporting participant recruitment and study implementation. We acknowledge the LLUH Institute for Community Partnerships Community Health Worker (CHW) Program for collaboration on CHW training, supervision, and intervention delivery. We are especially grateful to CHWs Rosy Perez Quintinalla and Candy Rangel for their compassionate, consistent, participant-centered support throughout the study. We also acknowledge the Smoke-Free HOPE Program Coordination Team for project management, recruitment, data collection, and operational support.

