## Supplementary material for "Pilot Feasibility Study of a Community Health Worker-delivered Tobacco Cessation Intervention During Pregnancy: The Smoke-Free HOPE Randomized Controlled Trial": Tables 1-4

**Table 1. Core Intervention Elements of the Smoke-Free HOPE Study Arms**

| **Core Element** | **CHW-Brief Advice (CHW-BA)** | **CHW-Patient Navigation (CHW-PN))** |
| --- | --- | --- |
| Primary mechanism | Direct behavioral support to enhance motivation, self-efficacy, quit planning, and relapse prevention | Navigation-based support to increase access to and engagement with evidence-based cessation resources |
| CHW Role | Deliver structured cessation counseling and reinforce behavioral skill development during weekly participant encounters | Facilitate identification, enrollment, and sustained participation in cessation programs and supportive resources |
| Cessation support strategy | Adapted the SCRIPT™-based brief advice model using motivational communication and pregnancy-tailored cessation education | Ask-Advise-Connect-informed navigation approach linking participants to quitlines, digital tools, and community cessation services |
| **Shared Components across Both Intervention Arms** | | |
| Session Format | Up to eight weekly sessions delivered through home visits or telehealth | |
| Motivational approach | Participant-centered motivational communication strategies | |
| Prenatal care integration | Coordination with prenatal care and referral for social needs support when indicated | |
| Supportive services | Screening and navigation for health-related social needs and community resources | |
| Participant incentives | Incentives provided for participation in study activities | |

**Table 2. Demographic Characteristics and Tobacco Product Use Among Smoke-Free HOPE Participants**

| **Table 2.** |  |  |  |  |  |  |  |  |
| --- | --- | --- | --- | --- | --- | --- | --- | --- |
| *Demographic Characteristics and Tobacco Product Use Among Smoke-Free HOPE Participants* | | | | | | | | |
| **Characteristic** | |  | **CHW-BA (n=17)** | **CHW-PN (n=15)** |  | **p-value** |  | **Total (n=32)** |
|  |  |  | **n (%)** | **n (%)** |  |  |  | **n (%)** |
| **Age** |  |  |  |  |  | 0.74^§^ |  |  |
|  | Mean (SD) [Range] |  | 30 (5.8) [20- 40] | 31 (7.3) [20-45] |  |  |  | 30.7 (6.5) [20-45] |
| **Annual Household Income** | |  |  |  |  | 0.92^§^ |  |  |
|  | Median [range] |  | $25,100 [$0- $75,000] | $20,000 [$0- $90,000] |  |  |  | $25,000 [$0, $90,000] |
|  | Missing/ "Don't Know" |  | 5 (16%) | 4 (13%) |  |  |  | 9 (28%) |
| **Sex at birth** |  |  |  |  |  | - |  |  |
|  | Female |  | 17 (100%) | 15 (100%) |  |  |  | 32 (100%) |
| **Sexual orientation** | |  |  |  |  | 0.60 |  |  |
|  | Straight, heterosexual |  | 13 (77%) | 14 (93%) |  |  |  | 27 (84%) |
|  | Lesbian/ Gay |  | 1 (6%) | 0 (0%) |  |  |  | 1 (3 %) |
|  | Bisexual/Pansexual |  | 3 (18 %) | 1 (7%) |  |  |  | 4 (13%) |
| **Race/Ethnicity** | |  |  |  |  | 0.91 |  |  |
|  | Latino/Hispanic |  | 4 (24%) | 3 (20%) |  |  |  | 7 (22%) |
|  | White |  | 1 (6%) | 2 (13%) |  |  |  | 3 (9%) |
|  | Black/African American |  | 8 (47%) | 6 (40%) |  |  |  | 14 (44%) |
|  | More than one race |  | 4 (24%) | 3 (20%) |  |  |  | 7 (22%) |
|  | Other |  | 0 (0%) | 1 (7%) |  |  |  | 1 (3%) |
| **Education** |  |  |  |  |  | 0.47 |  |  |
|  | Grade school or less |  | 0 (0%) | 1 (7%) |  |  |  | 1 (3%) |
|  | High school/ equivalent |  | 8 (47%) | 8 (53%) |  |  |  | 16 (50%) |
|  | College/University or higher |  | 1 (6%) | 1 (7%) |  |  |  | 2 (6%) |
|  | Community College |  | 3 (18%) | 0 (0%) |  |  |  | 3 (9%) |
|  | Vocational/Business/Trade School |  | 4 (24%) | 3 (20%) |  |  |  | 7 (22%) |
|  | Missing |  | 1 (6%) | 2 (13%) |  |  |  | 3 (9%) |
| **Employment** | |  |  |  |  | 0.54 |  |  |
|  | Full-time (> 30 hours/week) |  | 2 (12%) | 4 (27%) |  |  |  | 6 (19%) |
|  | Part-time (< 30 hours/week) |  | 3 (18%) | 2 (13%) |  |  |  | 5 (16%) |
|  | Unemployed |  | 12 (71%) | 8 (53%) |  |  |  | 20 (63%) |
|  | Missing |  | 0 (0%) | 1 (7%) |  |  |  | 1 (3%) |
| **Marital Status** | |  |  |  |  | 0.79 |  |  |
|  | Married |  | 2 (12%) | 5 (33%) |  |  |  | 7 (22%) |
|  | Living with partner |  | 7 (41%) | 5 (33%) |  |  |  | 12 (38%) |
|  | Separated |  | 1 (6%) | 0 (0%) |  |  |  | 1 (3%) |
|  | Widowed |  | 1 (6%) | 0 (0%) |  |  |  | 1 (3%) |
|  | Single, not married |  | 5 (29%) | 3 (20%) |  |  |  | 8 (25%) |
|  | Legally single, with partner |  | 1 (6%) | 1 (6%) |  |  |  | 2 (6%) |
|  | Missing |  | 0 (0%) | 1 (7%) |  |  |  | 1 (3%) |
| **Public Program Participation^†^** | |  |  |  |  |  |  |  |
|  | TANF/CalWORKS |  | 10 (59%) | 7 (47%) |  | 0.59 |  | 17 (53%) |
|  | CalFresh |  | 14 (82%) | 8 (53%) |  | 0.17 |  | 22 (69%) |
|  | Supplemental Security Income |  | 1 (6%) | 2 (13%) |  | 0.39 |  | 3 (9%) |
|  | WIC |  | 11 (65%) | 9 (60%) |  | 0.85 |  | 20 (63%) |
|  | Child Support |  | 1 (6%) | 1 (7%) |  | 0.73 |  | 2 (6%) |
| **Food Insecurity*** | |  |  |  |  | 0.85 |  |  |
|  | Food Insecure |  | 10 (59%) | 8 (53%) |  |  |  | 18 (56%) |
|  | Food Secure |  | 7 (41%) | 6 (40%) |  |  |  | 13 (41%) |
|  | Missing |  | 0 (0%) | 1 (7%) |  |  |  | 1 (3%) |
| **Access to Healthcare** | |  |  |  |  | 0.46 |  |  |
|  | Private / Purchased through Covered CA |  | 0 (0%) | 0 (0%) |  |  |  | 0 (0%) |
|  | Insurance through Employer (self/spouse) |  | 0 (0%) | 3 (20%) |  |  |  | 3 (9%) |
|  | Medi-CAL/Medicaid/ IEHP |  | 17 (100%) | 11 (73%) |  |  |  | 28 (88%) |
|  | Missing |  | 0 (0%) | 1 (7%) |  |  |  | 1 (3%) |
| **Tobacco Product Use** | |  |  |  |  |  |  |  |
| **Combustible Tobacco*** | |  |  |  |  |  |  |  |
|  | Cigarettes |  | 5 (29%) | 8 (53%) |  |  |  | 13 (41%) |
|  | Cigars/Blunts |  | 1 (6%) | 0 (0%) |  |  |  | 1 (3%) |
|  | Other^†^ |  | 0 (0%) | 0 (0%) |  |  |  | 0 (0%) |
|  | Electronic Nicotine Delivery Systems (ENDS)^§^* |  | 8 (47%) | 9 (60%) |  |  |  | 17 (53%) |
|  | Smokeless Tobacco^◊^ |  | 1 (6%) | 2 (14%) |  |  |  | 3 (9%) |
|  | Mixed use of tobacco products |  | 2 (12%) | 3 (20%) |  |  |  | 5 (16%) |
|  | Cannabis use (in past year) |  | 12 (71%) | 10 (67%) |  |  |  | 22 (69%) |
| **Substance Co-use** | |  |  |  |  |  |  |  |
|  | Tobacco and Alcohol** |  | 2 (12%) | 1 (7%) |  |  |  | 3 (9%) |
| *^§^ The p-value for age is from a two-sample t-test assuming equal variances, t(30) = -0.33, 95% CI for the difference [-5.5, 4.0]; the folded F test for equality of variances was non-significant (p = 0.38). The p-value for annual household income is from a two-sample t-test assuming equal variances, t(21) = -0.10, 95% CI for the difference [-22645, 20651]; the folded F test for equality of variances was non-significant (p = 0.68). The remaining p-values use Fisher's exact test for categorical variables because expected cell counts are too small for chi-square.*  *^†^ Count (n) and percentages do not sum to 100% as participants could be enrolled in more than one program.*  *^*^ Food insecurity was determined using the USDA 6-Item Scale with a raw score ≥ 2* | | | | | | | | |
| ** Used within the last 30 days*  *† Cigarillos, hookah/shisha pipes, pipes*  *§ Disposables, devices with replaceable pods, devices with refillable tanks, mod systems*  *◊ Participant has once used smokeless tobacco or nicotine products fairly regularly*  *** Alcohol usage defined by having drunk any alcohol in the past year (Audit-C score > 0)* | | | | | | | | |

**Table 3. Acceptability Survey Findings (n=16)**

| **Participant Satisfaction** | | | | | | | |
| --- | --- | --- | --- | --- | --- | --- | --- |
|  | **Overall program** | | **Unsatisfactory** | **Little Satisfaction** | **Somewhat Satisfied** | **Very Satisfied** |  |
|  |  |  | 0% | 0% | 9% | 91% |  |
|  | **Support Received** | | **Very Dissatisfied** | **Dissatisfied** | **Neutral** | **Satisfied** | **Very Satisfied** |
|  |  |  | 0% | 0% | 0% | 9% | 91% |
|  | **Likely to recommend SFH** | | **Definitely would not** | **Probably not** | **Unsure** | **Probably would** | **Definitely would** |
|  |  |  | 0% | 0% | 0% | 0% | 100% |
|  | **CHW Overall Communication Rating (1= very dissatisfied, 10= very satisfied)** | | | | | | |
|  |  | Mean, [Range] | 9.7, [7 - 10] \| |  |  |  |  |
| **Critical Factors** | | | | | | | |
|  | **1. Focus on my individual strengths** | | |  |  |  |  |
|  |  |  | **not at all/ poorly** | **a little/ slightly** | **moderate** | **very** | **absolutely essential** |
|  |  | Importance |  |  |  | 36% | 64% |
|  |  | Achieved |  |  | 9% | 18% | 73% |
|  | **2. Relationship to encourage change** | |  |  |  |  |  |
|  |  |  | **not at all/ poorly** | **a little/ slightly** | **moderate** | **very** | **absolutely essential** |
|  |  | Importance |  |  |  | 9% | 91% |
|  |  | Achieved |  |  | 9% | 18% | 73% |
|  | **3. Reliability and consistency** | |  |  |  |  |  |
|  |  |  | **not at all/ poorly** | **a little/ slightly** | **moderate** | **very** | **absolutely essential** |
|  |  | Importance |  |  |  | 9% | 91% |
|  |  | Achieved |  |  |  | 9% | 91% |
|  | **4. Connection to culture** | |  |  |  |  |  |
|  |  |  | **not at all/ poorly** | **a little/ slightly** | **moderate** | **very** | **absolutely essential** |
|  |  | Importance |  | 9% |  | 37% | 55% |
|  |  | Achieved | 9% | 18% |  | 18% | 55% |
|  | **5. Non-judgmental approach** | |  |  |  |  |  |
|  |  |  | **not at all/ poorly** | **a little/ slightly** | **moderate** | **very** | **absolutely essential** |
|  |  | Importance | 9% |  |  | 9% | 82% |
|  |  | Achieved |  |  |  | 10% | 90% |
|  | **6. Setting clear rules and boundaries** | | | | | | |
|  |  |  | **not at all/ poorly** | **a little/ slightly** | **moderate** | **very** | **absolutely essential** |
|  |  | Importance | 9% |  | 9% | 18% | 64% |
|  |  | Achieved |  |  |  | 10% | 90% |
|  | **7. Modeling openness, honesty, hope, and trust** | | |  |  |  |  |
|  |  |  | **not at all/ poorly** | **a little/ slightly** | **moderate** | **very** | **absolutely essential** |
|  |  | Importance |  |  |  | 18% | 82% |
|  |  | Achieved |  |  |  | 18% | 82% |
|  | **8. Feeling self-motivated, safe to try new things** | | |  |  |  |  |
|  |  |  | **not at all/ poorly** | **a little/ slightly** | **moderate** | **very** | **absolutely essential** |
|  |  | Importance |  |  |  | 36% | 64% |
|  |  | Achieved |  |  |  | 27% | 73% |
|  | **9. Celebrating small achievements and positive changes** | | | | | | |
|  |  |  | **not at all/ poorly** | **a little/ slightly** | **moderate** | **very** | **absolutely essential** |
|  |  | Importance |  |  |  | 9% | 91% |
|  |  | Achieved |  |  |  | 9% | 91% |
| **Perceived Usefulness and Support** | | | | | | | |
|  | ***Commit to Quit* Video (n=4)** | | **not helpful** | **a little helpful** | **somewhat helpful** | **very helpful** |  |
|  |  |  |  |  | 50% | 50% |  |
|  | ***A pregnant women's guide to quit smoking* (n=5)** | | **not helpful** | **a little helpful** | **somewhat helpful** | **very helpful** |  |
|  |  |  |  | 20% | 20% | 60% |  |
|  | **CHW resources (n=6)** | | **not helpful** | **a little helpful** | **somewhat helpful** | **very helpful** |  |
|  |  |  |  |  |  | 100% |  |
|  | **Kick-it California/Quitline resources (n=7)** | | **not helpful** | **a little helpful** | **somewhat helpful** | **very helpful** |  |
|  |  |  |  |  |  | 100% |  |

**Table 4. Joint Display of Quantitative and Qualitative Findings on Engagement, Acceptability, and Intervention Refinement**

| **Integration domain** | **Quantitative finding** | **Participant qualitative finding** | **Integrated interpretation** |
| --- | --- | --- | --- |
| **Engagement and acceptability** | 87% initiated CHW sessions and 66% completed all eight sessions. Participant satisfaction and CHW communication ratings were high. | Interviewed completers described CHWs as trusted, flexible, accessible, accountable, and nonjudgmental. Participants valued continued support through setbacks and competing demands. | **Convergence and complementarity:** Interviewed completers highly valued the relational and practical features of CHW support and perceived them as supporting continued engagement. |
| **Motivation, cessation challenges, and intervention fit** | 75% set a quit date, while 50% achieved 30-day point-prevalence abstinence from nicotine; 69% reported past-year cannabis use. | Pregnancy strengthened motivation, but participants described cessation as nonlinear and complicated by stress, cravings, and withdrawal, vaping accessibility, and social influences. In less common accounts, participants described cannabis or alcohol use as influencing their tobacco or e-cigarette use. Participants valued support that normalized setbacks and recognized gradual progress. | **Complementarity:** Strong motivation did not uniformly translate into abstinence. Participant accounts contextualized cessation experiences and informed priorities for vaping-specific cessation support and postpartum relapse prevention without establishing causes of cessation outcomes. |
| **Arm-specific resource use** | Survey respondents who evaluated intervention resources rated them highly. | In the brief-advice arm, the SCRIPT booklet was useful during CHW-guided sessions but was not consistently retained or revisited between visits. In the patient-navigation arm, CHWs helped participants overcome practical and technological barriers, facilitated contact with cessation counselors, and provided follow-up and accountability. | **Expansion:** Favorable resource ratings did not necessarily indicate sustained independent use. Participant accounts suggest that resource format, accessibility, and active CHW navigation influenced how participants accessed and used cessation resources. |

Note: Post-program interviews were conducted with intervention completers and were not designed to explain recruitment, nonparticipation, or disengagement. Accordingly, the joint display does not include recruitment and retention outcomes without corresponding qualitative dat
