## Supplementary material for "Pilot Feasibility Study of a Community Health Worker-delivered Tobacco Cessation Intervention During Pregnancy: The Smoke-Free HOPE Randomized Controlled Trial": Figure 1

CONSORT

TRANSPARENT REPORTING of TRIALS

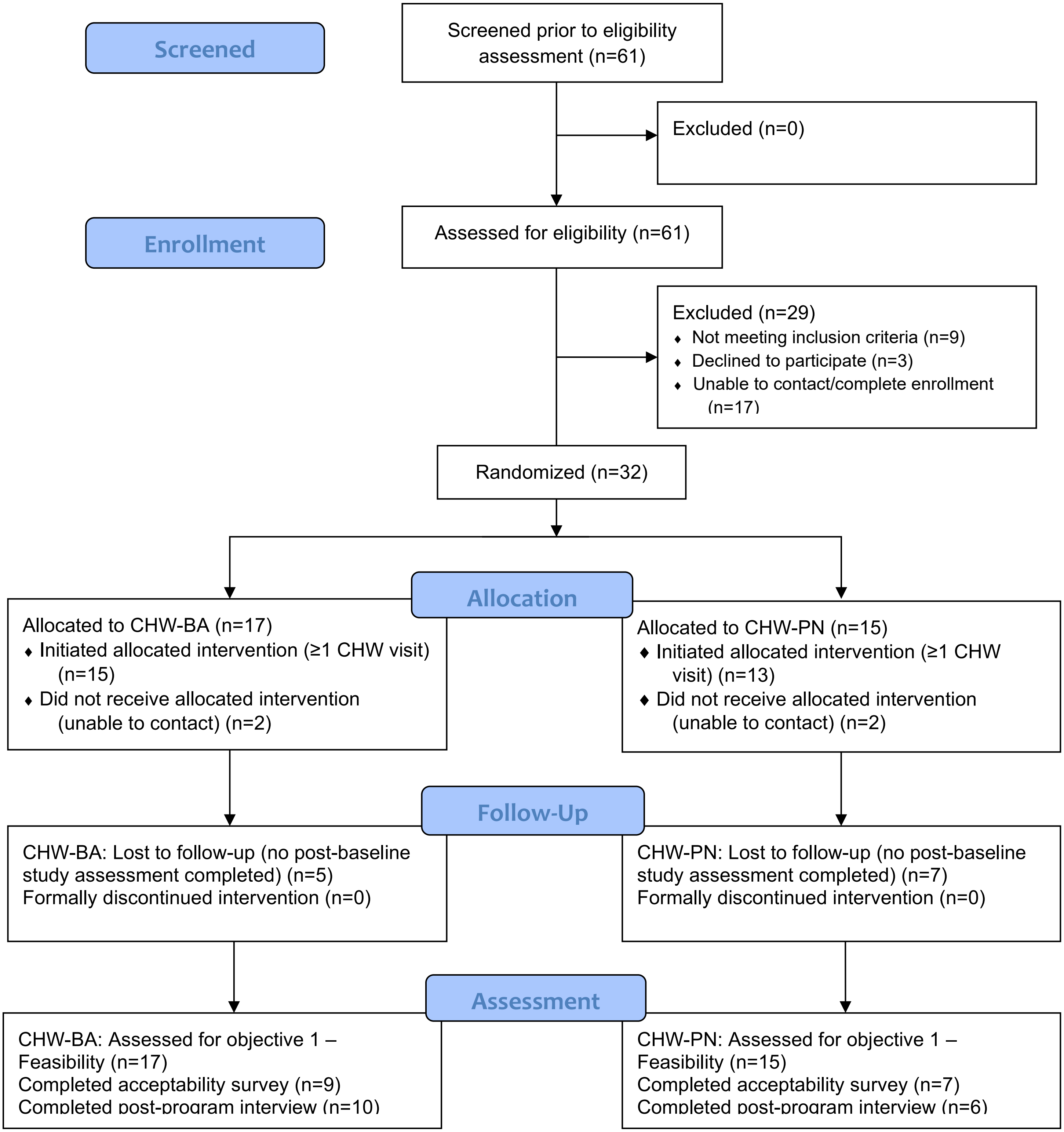


Note: The acceptability survey and the post-program qualitative interview samples each included 16 participants, but they were not identical.

Citation: Eldridge SM, Chan CL, Campbell MJ, Bond CM, Hopewell S, Thabane L, et al. CONSORT 2010 statement: extension to randomised pilot and feasibility trials. BMJ. 2016;355.
