## Supplemental File 2. Interview Guide for "Pilot Feasibility Study of a Community Health Worker-delivered Tobacco Cessation Intervention During Pregnancy: The Smoke-Free HOPE Randomized Controlled Trial"

### Post-Program Interview Guide [Participants]

Participant ID: ____________
Intervention Arm: ____________
Interview Date: ______________

---

**Introduction, Ground Rules, and Instructions**

Thank you for agreeing to participate in this interview. Your insights and experiences are invaluable in helping us understand how maternal smoking cessation programs support pregnant individuals and how quitting experiences may differ between combustible tobacco users and those who vape nicotine products.

The interview will last approximately 60 minutes. I will ask questions about your experiences with the Smoke-Free HOPE program, including what worked well, what could be improved, and any challenges you faced.

We will also ask about your experiences trying to quit smoking cigarettes and/or vaping to better understand differences in quitting experiences.

You may skip any question or stop the interview at any time.

---

**Ground Rules**

- **Confidentiality:** Your responses will be kept confidential and de-identified.
- **Voluntary Participation:** You may skip any question or stop at any time.
- **Respectful Dialogue:** No right or wrong answers.
- **Time and Pace:** Take breaks if needed.
- **Privacy:** Share only what you are comfortable sharing.
- **Recording:** Interviews may be recorded with your permission.

---

**Purpose**

This interview aims to gather feedback on your experiences with the Smoke-Free HOPE program and to explore differences in quitting experiences between combustible tobacco users and individuals who vape nicotine products or devices.

---

**Ice breaker**

#### I. Cessation Experiences: Tobacco and Vaping Use, Perceptions, and Quitting

##### Tobacco and Nicotine Use History

- Can you tell me about your history with tobacco or nicotine use? (Probe: How did you first get started?)

- What types of products have you used? (e.g., cigarettes, cigars, vaping/e-cigarettes, smokeless tobacco)
- When did you first start using each product?
- Are you currently using any of them?

**First Use of Vaping Products (if applicable)**

- Could you tell me about the first time you vaped? (Probe: How old were you? What was going on in your life at the time?)
    - What did you vape (nicotine, cannabis, flavors)?
    - What was it like?
    - What was the best or worst part?
    - Who were you with?

**Recent Use of Vaping Products (if applicable)**

- Think back to before and during the program: How did you vape (settings, frequency, alone or with others)?
    - Where do you normally vape?
    - Do you ever vape alone?
    - Did you use flavored nicotine or cannabis?
- Did your vaping change during the program? If so, how? (Reduced sessions? Changed products?)

**Perceived Benefits and Harms of Vaping or Smoking**

- Have you noticed any impact of vaping or smoking tobacco on your health?
    - How beneficial did you find it to vape or smoke? (e.g., morning sickness, anxiety, sleep, appetite)
    - What concerns or worries did you have about the harm of vaping during pregnancy?
- How did vaping or smoking affect your relationships with others? (Probe: Encouragement or discouragement?)
    - Stigma/judgment: How did you deal with judgment from others?

**Co-Use and Product-Specific Patterns**

- Are you currently using more than one type of tobacco or nicotine product or cannabis?
- If yes, which ones and how often?
- In what situations or settings do you tend to use each product?
- Do you use different products for different purposes (stress, social settings, cravings)?
- Do you consider one product more or less harmful than another? Why?
- Have you used any nicotine replacement therapy or quit-aid medications? Tell me more about your experience.

**Experiences Trying to Quit**

- Have you ever tried to quit smoking cigarettes? What motivated you? How many times have you tried? What helped or made it hard?
    - How would you describe the emotional or physical experience when trying to quit smoking? (Probe: cravings, stress, mood changes?)
- Have you ever tried to quit vaping? What led you to try? How did that compare to quitting cigarettes?
    - Were the challenges or triggers the same or different compared to quitting cigarettes?
    - What, if anything, helped you quit or reduce vaping? (Probe: specific people, programs, apps, strategies?)

**Comparison of Quitting Smoking vs. Quitting Vaping**

- How would you compare your experience trying to quit smoking versus trying to quit vaping?
    - Which did you find more difficult, and why? (Probe: physical cravings, social settings, stress, etc.)
    - Were your motivations or expectations different for each?

**Switching Between Products**

- Have you ever switched from cigarettes to vaping, or from vaping to cigarettes? What led to the switch?
- Did switching help reduce or change your tobacco/nicotine use?
- What was your experience like during that transition? Did it feel like progress toward quitting?

**Reflections and Support Needs**

- In your view, what makes quitting vaping different from quitting cigarettes?
- What types of messages, programs, or resources would be helpful for quitting either or both?
- Is there anything else you want to share about your experiences with tobacco or nicotine products?

---

#### II. Smoke-Free HOPE Program Feedback

**Initial Expectations and Perceptions**

- Before you participated in the Smoke-Free HOPE program, what were your expectations?
    - What did you hope to get out of it?
    - What was your felt need for support with tobacco or nicotine cessation?
- How did you first hear about the program?
- What motivated you to join?
    - What got you to say, "I'm interested"?
    - What got you to send the email inquiry?
    - why do they think patients are not responding to the Mychart?
    - Why do they think providers are not referring to our program?
- Did your feelings about the program change as you participated?
    - What was different from your expectations?
    - Were there any surprises?

**Study Recruitment and Enrollment Process**

- How did you hear about our program? [Probe based on their response]
    - **MyChart Invitation**:
        - What were your impressions of the invitation?
        - What motivated you to respond/contact us?
        - Did you have any hesitations about signing up? If so, what were they?
    - **Provider Recommendation/Referral**:
        - Which provider recommended the program (e.g., social worker, nurse, doctor)?
        - What motivated you to respond/contact us?
        - Did you have any hesitations about signing up? If so, what were they?
    - **Flyer**:
        - What were your first impressions of the flyer?
        - What motivated you to respond/contact us?
        - Did you have any hesitations about signing up? If so, what were they?
    - **Website**:
        - How did you find the Smoke-Free HOPE website?
        - What was your experience with navigating the website?
        - What motivated you to respond/contact us?
        - Did you have any hesitations about signing up? If so, what were they?
    - Other
- **What were the main factors that made you interested in joining the program? (Perceived benefit)**
    - Was it the cessation support, the incentives, or something else?
- Enrollment Process:
    - **How would you describe your experience with the enrollment process?**
        - Were the steps clear (e.g., phone screening, intake surveys)?
        - What could have made the process easier or clearer?

**Program Activities and Support**

- Which activities were most helpful (incentives, CHW visits, SCRIPT, Kick It CA, etc.)?
- Greatest impacts?
- Were there resources that helped you quit or stay quit?
- Least helpful parts?
- Any unexpected challenges, and how did you overcome them?

**Comfort and Communication with Community Health Worker (CHW)**

- How comfortable were you discussing tobacco/nicotine use with your CHW?
    - Did you feel heard?
    - Was anything challenging?
- How would you rate your CHW experience (1–5)? Why?
- Most helpful/least helpful parts of CHW support?
- Level of trust
- What do you think made the interaction with the CHW so effective, helpful, etc.?
- How much interaction did the CHW have with the rest of your household?
- What other types of support beyond the tobacco/nicotine use did the CHW offer?
    - What was most helpful?

**Study Materials and Resources**

- Impressions of program materials (e.g., SCRIPT booklet, surveys)?
- Did you use Kick It CA quit line or other resources?
    - If so, what was your experience?
    - Rate the resources (1–5)?
- Did you use any community-based resources?
    - What was your impression of those?

**Program Format and Length**

- Preference for in-person vs. telehealth? Why?
- Do you prefer any particular platform (FaceTime, WhatsApp, Zoom, MyChart)?
- How would you rate the program length and session frequency?

**Motivation and Engagement**

- What kept you engaged?
- What made participation difficult (barriers)?
- What strategies were most helpful for changing your behavior?
- What barriers still exist?
- Outlook: How confident are you in quitting tobacco/vaping?
    - What makes you feel confident or unsure?

**Benefits and Takeaways**

- What was the greatest impact of the program?
- Any unexpected benefits?
- Did it also benefit people around you?
- Any challenges resulting from program participation?
- Are there other things you wished the program would take into consideration?
    - Alcohol use?
    - Cannabis use?
    - Other coping mechanisms/strategies?

**Relevance for the Community**

- Would you recommend this program to others? Why or why not?
  - Friends? Providers?

**Suggestions for Improvement**

- What improvements would you recommend for future participants?
- Would you like to see this program integrated into routine prenatal care?
- If we were to offer group sessions led by CHWs, how interested would you be in participating?

**Closing Thoughts**

- Any final reflections?
- Anything else you would like to share?

---

**Final Notes:**

- Use follow-up probes when needed:
  - "Can you tell me more about that?"
  - "How did that make you feel?"
  - "Could you give me an example?"
  - "What did that look like for you?"
- Listen actively and allow participants to guide the conversation flow.

---
