## Supplemental File 3. Coding Tree for "Pilot Feasibility Study of a Community Health Worker-delivered Tobacco Cessation Intervention During Pregnancy: The Smoke-Free HOPE Randomized Controlled Trial"

### Qualitative Coding Framework and Analytic Approach

*Smoke-Free HOPE: A Pilot Feasibility Trial of Community Health Worker Models for Tobacco Cessation in Pregnancy*

The qualitative coding framework comprised seven primary domains and associated codes developed by deductive and inductive analytic processes. A cross-cutting analytic lens was subsequently applied to the coded material to identify implications for program refinement, implementation, and future delivery. Because this lens involved re-examining existing coded material rather than creating a separate coding category, it is presented separately from the primary coding framework.

#### Coding Framework Overview

**Domain 1. Tobacco and Nicotine Use History**
- Convenience and Accessibility of Vaping
- Behavioral and Sensory Dependence

**Domain 2. Perceptions of Harm and Risk**
- Vaping Perceived as Safer

**Domain 3. Motivation and Readiness to Quit**
- Pregnancy and Infant Health Motivation

**Domain 4. Withdrawal, Relapse, and Emotional Coping**
- Withdrawal Symptoms
- Emotional Coping and Stress Management
- Trauma, Grief, and Emotional Escape

**Domain 5. Social and Environmental Context**
- Peer and Social Influence
- Shame, Concealment, and Stigma

**Domain 6. CHW Relationship and Program Acceptability**
- CHW Emotional Support

**Domain 7. Engagement and Retention**
- Program Flexibility and Accessibility

#### Full Coding Tree

| Domain (Purpose) | Code | Definition |
|---|---|---|
| **Domain 1: Tobacco and Nicotine Use History**<br>*To understand initiation pathways, progression of nicotine dependence, transitions between products, and contextual patterns of use.* | **Convenience and Accessibility of Vaping** | Participant describes vaping as easier to use, conceal, or access than cigarettes. |
| | **Behavioral and Sensory Dependence** | Participant describes habitual, automatic, or sensory-driven use patterns (e.g., hand-to-mouth behavior, device proximity, flavor/aesthetic attraction) that may operate independently of nicotine content. |
| **Domain 2: Perceptions of Harm and Risk**<br>*To examine participant beliefs regarding nicotine products, vaping safety, pregnancy-related risk, and perceived health consequences.* | **Vaping Perceived as Safer** | Participant describes vaping as healthier, cleaner, safer, or less harmful than cigarettes. |
| **Domain 3: Motivation and Readiness to Quit**<br>*To identify factors influencing cessation attempts and readiness for behavior change.* | **Pregnancy and Infant Health Motivation** | Participant identifies pregnancy, fetal health, infant wellbeing, or parenting identity as motivation for cessation. |

| Domain (Purpose) | Code | Definition |
|---|---|---|
| **Domain 4: Withdrawal, Relapse, and Emotional Coping**<br>*To understand emotional and physical dimensions of cessation attempts.* | **Withdrawal Symptoms** | Participant describes emotional or physical withdrawal experiences during cessation attempts. |
| | **Emotional Coping and Stress Management** | Participant describes nicotine use as a coping strategy for emotional distress, anxiety, or stress. |
| | **Trauma, Grief, and Emotional Escape** | Participant describes nicotine use in the context of bereavement, acute grief, accumulated stress, or emotional avoidance/escape. |
| **Domain 5: Social and Environmental Context**<br>*To identify external influences shaping nicotine use and cessation outcomes.* | **Peer and Social Influence** | Participant describes peers, partners, family, or social settings influencing nicotine use or cessation. |
| | **Shame, Concealment, and Stigma** | Participant describes hiding tobacco/nicotine use from others, pregnancy-related guilt, fear of judgment, or healthcare-related stigma. |
| **Domain 6: CHW Relationship and Program Acceptability**<br>*To assess participant experiences with CHW-delivered intervention components.* | **CHW Emotional Support** | Participant describes emotional support, validation, or encouragement provided by CHWs. |
| **Domain 7: Engagement and Retention**<br>*To identify facilitators and barriers affecting participation and sustained involvement.* | **Program Flexibility and Accessibility** | Participant describes intervention flexibility, accessibility, or participant-centered communication. |

**Cross-cutting Analytic Lens: Program Refinement and Implementation Insights**

| Analytic Lens | Application |
|---|---|
| Program Refinement and Implementation Insights | Previously coded material across Domains 1–7 was revisited to identify implications for program refinement, implementation, and future delivery. |
