## Supplemental File 4. SDOH needs reported for "Pilot Feasibility Study of a Community Health Worker-delivered Tobacco Cessation Intervention During Pregnancy: The Smoke-Free HOPE Randomized Controlled Trial"

**Supplemental Table 4. Social Determinants of Health (SDOH)-related needs reported during CHW sessions, by intervention arm and overall (n=28)**

| | Brief Advice (n=15)* | | Patient Navigation (n=13) | | Combined | |
|---|---|---|---|---|---|---|
| **SDOH** | **n** | **%** | **n** | **%** | **n** | **%** |
| Food Insecurity | 8 | 57% | 11 | 85% | 19 | 70% |
| Lack of Employment | 7 | 50% | 5 | 39% | 12 | 44% |
| Housing Insecurity | 6 | 43% | 10 | 77% | 16 | 59% |
| Life Skills | 4 | 29% | 12 | 92% | 16 | 59% |
| Victim Services | 3 | 21% | 1 | 8% | 4 | 15% |
| Legal Issues | 2 | 14% | 6 | 46% | 8 | 30% |
| Substances or Alcohol Dependence | 1 | 7% | 2 | 15% | 3 | 11% |
| Inadequate physical health | 1 | 7% | 1 | 8% | 2 | 7% |
| Poverty | 1 | 7% | 5 | 39% | 6 | 22% |
| Lack of educational opportunities | 1 | 7% | 5 | 39% | 6 | 22% |
| Inadequate mental health | 0 | 0% | 2 | 15% | 2 | 7% |

*There are a total of n=15 in CHW-BA; however, because one participant in the CHW-BA intervention arm had missing SDOH data across all CHW surveys, n=14 was used to calculate the % of each SDOH in the CHW_BA group.
The count and % do not sum up to 100% because SDOH are not mutually exclusive; participants can identify multiple at a given time.

Supplemental Figure 4. Overall prevalence of SDOH-related needs reported during CHW sessions

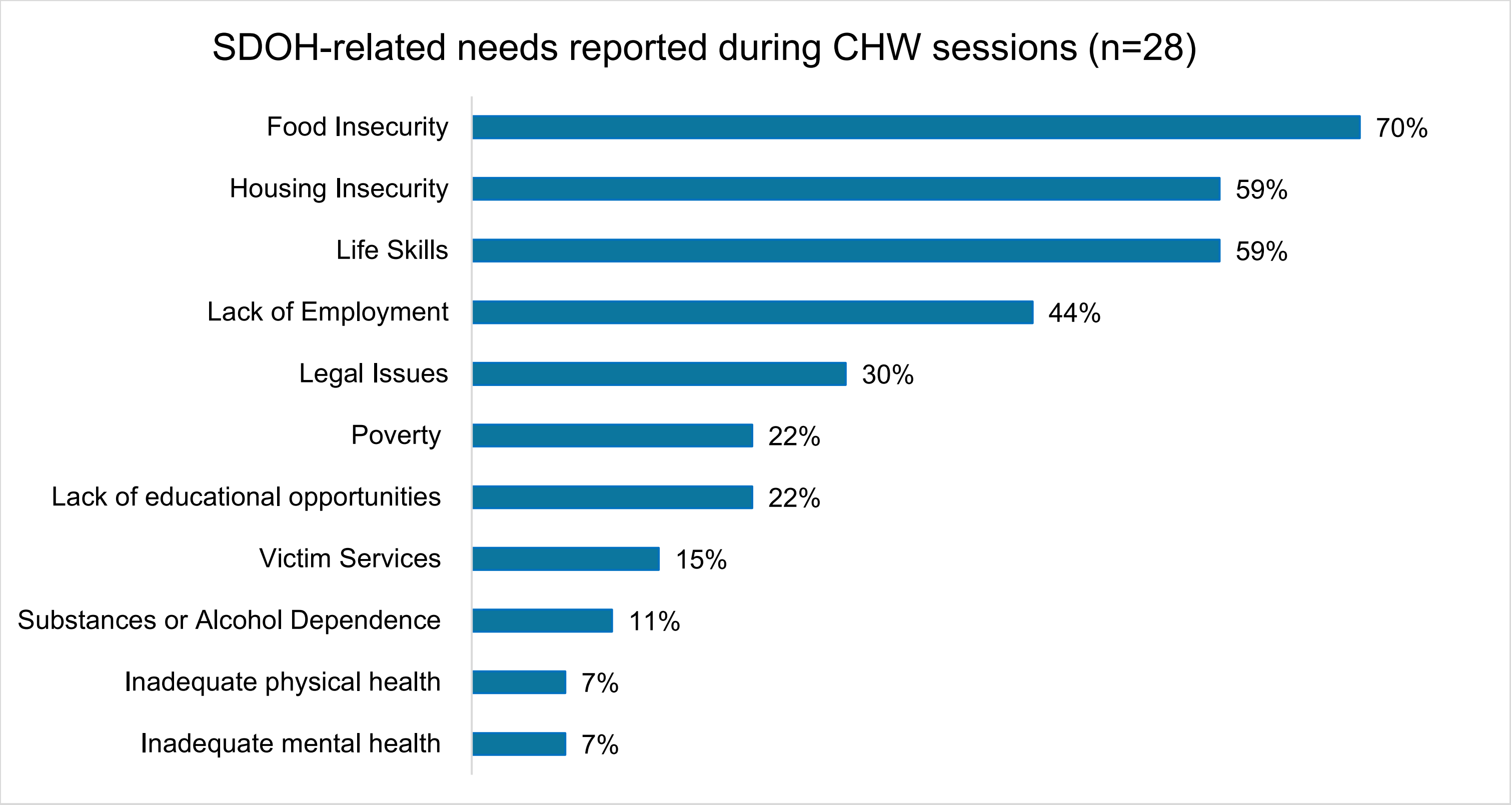
